# Determinants of penetrance and variable expressivity in monogenic metabolic conditions across 77,184 exomes

**DOI:** 10.1101/2020.09.22.20195529

**Authors:** Julia Goodrich, Moriel Singer-Berk, Rachel Son, Abigail Sveden, Jordan Wood, Eleina England, Joanne B. Cole, Ben Weisburd, Nick Watts, Zachary Zappala, Haichen Zhang, Kristin A. Maloney, Andy Dahl, Carlos A. Aguilar-Salinas, Gil Atzmon, Francisco Barajas-Olmos, Nir Barzilai, John Blangero, Eric Boerwinkle, Lori L. Bonnycastle, Erwin Bottinger, Donald W Bowden, Federico Centeno-Cruz, John C. Chambers, Nathalie Chami, Edmund Chan, Juliana Chan, Ching-Yu Cheng, Yoon Shin Cho, Cecilia Contreras-Cubas, Emilio Córdova, Adolfo Correa, Ralph A. DeFronzo, Ravindranath Duggirala, Josée Dupuis, Ma. Eugenia Garay-Sevilla, Humberto García-Ortiz, Christian Gieger, Benjamin Glaser, Clicerio González-Villalpando, Ma Elena Gonzalez, Niels Grarup, Leif Groop, Myron Gross, Christopher Haiman, Sohee Han, Craig L Hanis, Torben Hansen, Nancy L. Heard-Costa, Brian E Henderson, Juan Manuel Malacara Hernandez, Mi Yeong Hwang, Sergio Islas-Andrade, Marit E Jørgensen, Hyun Min Kang, Bong-Jo Kim, Young Jin Kim, Heikki A. Koistinen, Jaspal Singh Kooner, Johanna Kuusisto, Soo-Heon Kwak, Markku Laakso, Leslie Lange, Jong-Young Lee, Juyoung Lee, Donna M. Lehman, Allan Linneberg, Jianjun Liu, Ruth J.F. Loos, Valeriya Lyssenko, Ronald C. W. Ma, Angélica Martínez-Hernández, James B. Meigs, Thomas Meitinger, Elvia Mendoza-Caamal, Karen L. Mohlke, Andrew D. Morris, Alanna C. Morrison, Maggie CY Ng, Peter M. Nilsson, Christopher J. O’Donnell, Lorena Orozco, Colin N. A. Palmer, Kyong Soo Park, Wendy S. Post, Oluf Pedersen, Michael Preuss, Bruce M. Psaty, Alexander P. Reiner, Cristina Revilla-Monsalve, Stephen S Rich, Jerome I Rotter, Danish Saleheen, Claudia Schurmann, Xueling Sim, Rob Sladek, Kerrin S Small, Wing Yee So, Xavier Soberón, Timothy D Spector, Konstantin Strauch, Tim M Strom, E Shyong Tai, Claudia H.T. Tam, Yik Ying Teo, Farook Thameem, Brian Tomlinson, Russell P. Tracy, Tiinamaija Tuomi, Jaakko Tuomilehto, Teresa Tusié-Luna, Rob M. van Dam, Ramachandran S. Vasan, James G Wilson, Daniel R Witte, Tien-Yin Wong, Lizz Caulkins, Noël P. Burtt, Noah Zaitlen, Mark I. McCarthy, Michael Boehnke, Toni I. Pollin, Jason Flannick, Josep M. Mercader, Anne O’Donnell-Luria, Samantha Baxter, Jose C. Florez, Daniel MacArthur, Miriam S. Udler-Aubrey, for AMP-T2D-GENES Consortia

## Abstract

Hundreds of thousands of genetic variants have been reported to cause severe monogenic diseases, but the probability that a variant carrier will develop the disease (termed penetrance) is unknown for virtually all of them. Additionally, the clinical utility of common polygenetic variation remains uncertain. Using exome sequencing from 77,184 adult individuals (38,618 multi-ancestral individuals from a type 2 diabetes case-control study and 38,566 participants from the UK Biobank, for whom genotype array data were also available), we applied clinical standard-of-care gene variant curation for eight monogenic metabolic conditions. Rare variants causing monogenic diabetes and dyslipidemias displayed effect sizes significantly larger than the top 1% of the corresponding polygenic scores. Nevertheless, penetrance estimates for monogenic variant carriers averaged below 60% in both studies for all conditions except monogenic diabetes. We assessed additional epidemiologic and genetic factors contributing to risk prediction, demonstrating that inclusion of common polygenic variation significantly improved biomarker estimation for two monogenic dyslipidemias.

## Introduction

Healthcare providers and researchers are increasingly faced with interpreting genetic sequence data collected from individuals who are asymptomatic or for whom limited clinical information is available. Standard clinical practice for reporting whole exome and genome sequencing results may involve risk assessment for genetic variation causing conditions of known relevance to the individual and also potentially impactful variants unrelated to the primary indication for testing (termed “secondary genetic findings,” for example the American College of Medical Genetics and Genomics (ACMG) list of 59 medically actionable genes^1-4^). Thus, predicting the risk conferred by genetic findings in individuals who are not known to have the relevant conditions is of critical importance, but remains a challenge^5^. Furthermore, the scope of genetic variation interpreted in current clinical genetics practice is predominantly limited to rare monogenic “Mendelian” disease variants with large predicted effect sizes, leaving the vast majority of the genome, including common variants, unassessed. Recent studies have suggested that a high burden of common genetic variation may confer increased disease risk equivalent in magnitude to carrying rare monogenic variants^6^; however, this equivalency has recently been called into question^7^, and it remains uncertain whether and how to integrate polygenic scores capturing common genetic variation into medical care^8^.

Clinical application of genomic sequence data requires identification of medically significant genetic variants and estimation of their impact. In recent years, detailed guidelines from the ACMG and the Association for Molecular Pathology (AMP)^9^ have provided standards for reporting clinically significant variants which have been implemented by approximately 95% of clinical laboratories internationally^10^. However, the probability that carriers of such variants will manifest the given condition (termed “penetrance”) is unknown or uncertain for the vast majority of reported pathogenic variants^5^. Furthermore, individuals with the same genotype may exhibit variable degrees of phenotype expression (termed “variable expressivity”)^11,12^. Estimates of penetrance and expressivity traditionally have been derived from studies focusing on individuals with a given condition and their family members; this approach suffers from ascertainment bias, since the proband, who came to clinical attention due to having the condition, may share other genetic and/or environmental factors influencing manifestation of the condition with their family members^12,13^. Interpretation of rare variants identified by sequencing is further complicated by limited or no data available from any source, including families, to assess penetrance^5^.

Large-scale population-based and cohort studies with both sequence and phenotype data offer an opportunity to estimate penetrance and expressivity with less upward bias compared to family or case-control studies. In fact, population-based studies may have a healthy-participant bias which could provide downwardly biased estimates of penetrance^14^. Recent studies attempting to connect large-scale genetic and phenotypic data have noted reduced penetrance estimates compared to those previously reported; however, these recent studies were limited by sample size and/or application of less stringent curation of genetic variants than the current clinical standard of care ACMG/AMP guideline approach^7,14-17^. Additionally, further characterization of additional epidemiologic and genetic factors, such as phenotypic ascertainment and polygenic risk, is needed for accurate prediction of penetrance and expressivity for rare monogenic variants.

Here we present analyses performed in two separate datasets: 38,618 exomes from individuals ascertained as part of multi-ancestral type 2 diabetes (T2D) case-control studies, and 38,566 exomes from individual volunteers in the UK Biobank (UKB). Our analyses focused on traits with complex genetic architectures, involving rare and common genetic contribution, and well-defined biomarkers. These included diabetes (maturity-onset diabetes of the young (MODY), neonatal diabetes, autosomal dominant lipodystrophy) and disorders of LDL cholesterol, HDL cholesterol, triglycerides, and obesity. In addition to performing stringent curation using the ACMG/AMP criteria^9^ to generate a set of clinically significant genetic variants, we have also calculated polygenic scores in the UKB dataset to assess the cumulative impact of common variation on the same phenotypes. These data allow us to make a direct comparison between monogenic and polygenic risk, and to assess the contribution of polygenic risk to expressivity for carriers of monogenic variants.

## Results

### Identification of high confidence clinically significant variants enhances risk stratification

We studied two distinct datasets for which both individual-level exome sequence and phenotypic data were available (N=77,184): a compilation of multi-ancestral case control-studies for T2D, involving 22,875 T2D (or prediabetes) cases (see **Methods**) and 15,743 controls from the T2D-GENES and AMP-T2D consortia^18^, (referred to subsequently as AMP-T2D-GENES); and 38,566 unrelated individuals of European origin from the UKB^19^ (see **Methods, Supplementary Table 1**). Our analyses focused on 26 genes offered by clinical laboratories the United States for evaluation of monogenic forms of diabetes or diabetes-related traits through autosomal dominant modes of inheritance: MODY most commonly offered in panel testing (*GCK, HNF1A, HNF1B, HNF4A, PDX1*), an extended set of purported MODY genes less frequently offered in panel testing (*AKT2, KLF11, APPL1, ABCC8, KCNJ11, NEUROD1, CEL, INS*), neonatal diabetes (*ABCC8, GATA4, GATA6, HNF1B, INS, KCNJ11*), lipodystrophy (*AKT2, LMNA, PLIN1, PPARG*), elevated LDL cholesterol (*LDLR, APOB*), low serum LDL cholesterol (*APOB, PCSK9*), elevated serum HDL cholesterol (*CETP*), hypertriglyceridemia (*APOA5, LPL*), and monogenic obesity (*MC4R*).

We performed stringent variant curation using the clinical gold standard ACMG/AMP criteria, blinded to carrier phenotypic data for two classes of variants: 276 variants previously reported to be clinically significant (designated “pathogenic” or “likely pathogenic”) in the ClinVar database^20^ or designated as disease-causing in review articles^21-23^; and 218 predicted loss of function (pLoF) variants in genes with supported loss-of-function mechanism of action (see **Methods**). Our approach was intended to capture high-confidence clinically significant variants, although notably excluded missense variants beyond those in the ClinVar database because of the low prior probability of disease relevance and the challenges of inferring pathogenicity for this variant class. In total across the AMP-T2D-GENES and UKB study exomes, 238 variants, representing 51% of all 463 variants curated, were determined by ACMG/AMP criteria to be clinically significant and were found in 626 carriers (**Figure 1, Supplementary Table 3, Supplementary Table 12**). Across the conditions, the clinically significant variants were observed in all represented ancestral groups (**Supplementary Figure 1**).

**Figure 1.**
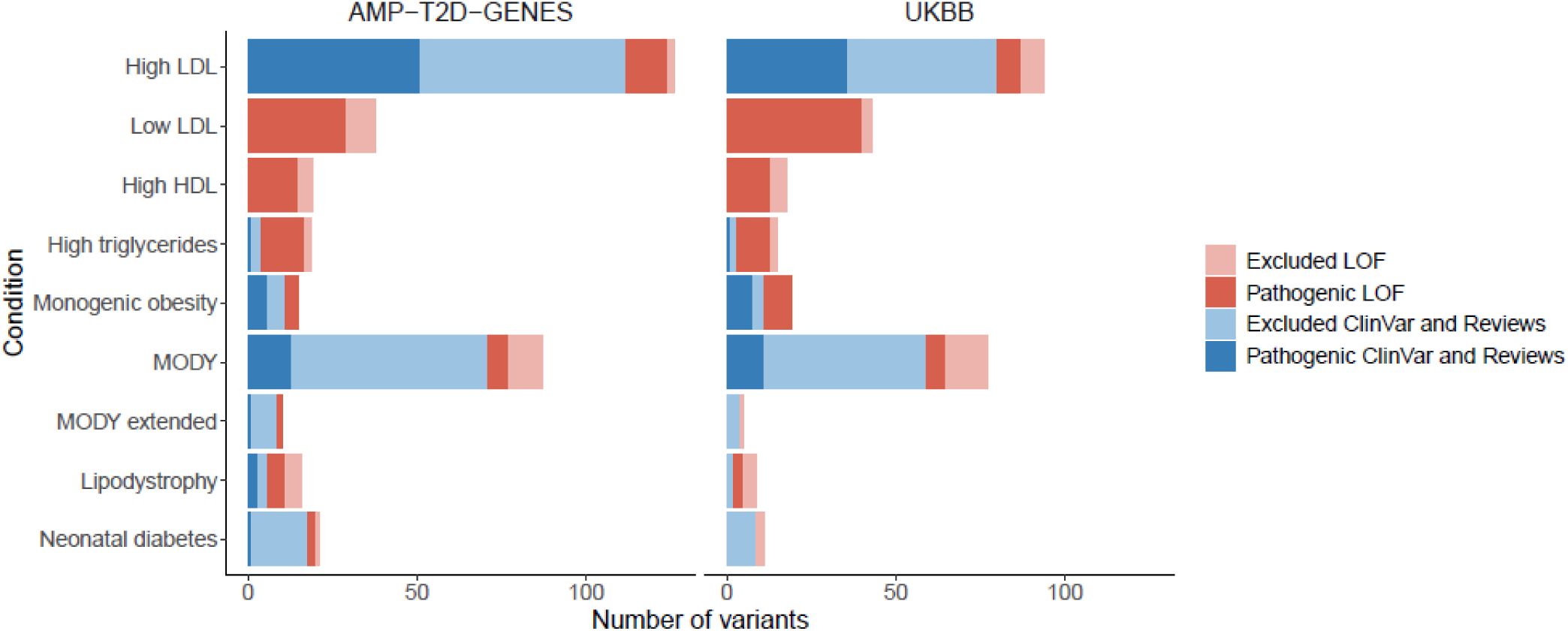
Curation of ClinVar and pLoF variants across the monogenic conditions. Total number of curated ClinVar/Review (blue) and pLoF (red) variants with carriers in AMP-T2D-GENES (left panel) and UKB (right panel). Darker color shades indicate variants determined to be clinically significant (pathogenic, likely pathogenic, or pLoF) and lighter shades indicate variants excluded during curation from further analysis.

We next assessed the impact of clinically significant monogenic variants on corresponding biomarkers, restricting analyses to conditions with at least ten carriers of variants in relevant genes (**Supplementary Table 3**). Monogenic variant carriers for dyslipidemias had significantly more extreme lipid trait values compared to non-carriers, with shifts of ∼55 mg/dL for both high and low LDL cholesterol conditions, ∼130 mg/dL for high triglycerides, and ∼16 md/dL for high HDL cholesterol (*P*<10^−5^ for all; adjusted for age, sex, and 10 PCs; **Table 1**). For monogenic obesity, *MC4R* variant carriers had ∼2 kg/m^2^ higher mean body mass index (BMI) than non-carriers in both datasets, however this difference reached significance only in UKB (*P*=0.063 AMP-T2D-GENES, *P*=0.006 UKB). Despite differences in the study populations and designs in AMP-T2D-GENES and UKB, the effect sizes of clinically significant variants on relevant biomarkers were remarkably consistent across the two studies for dyslipidemia and obesity gene sets, once the former was adjusted for lipid medication use (**Table 1, Supplementary Table 4**). MODY variant carriers had significantly increased odds of having diabetes compared to non-carriers in both studies (OR>7, *P*<10^−4^; **Table 1, Supplementary Table 4**); differences in risk estimates between the two studies were likely influenced by ascertainment practices in AMP-T2D-GENES, as it was a T2D case-control study and several sub-studies intentionally excluded diabetes cases with clinical features suggestive of MODY^18^ (**Supplementary Table 2**).

**Table 1:**
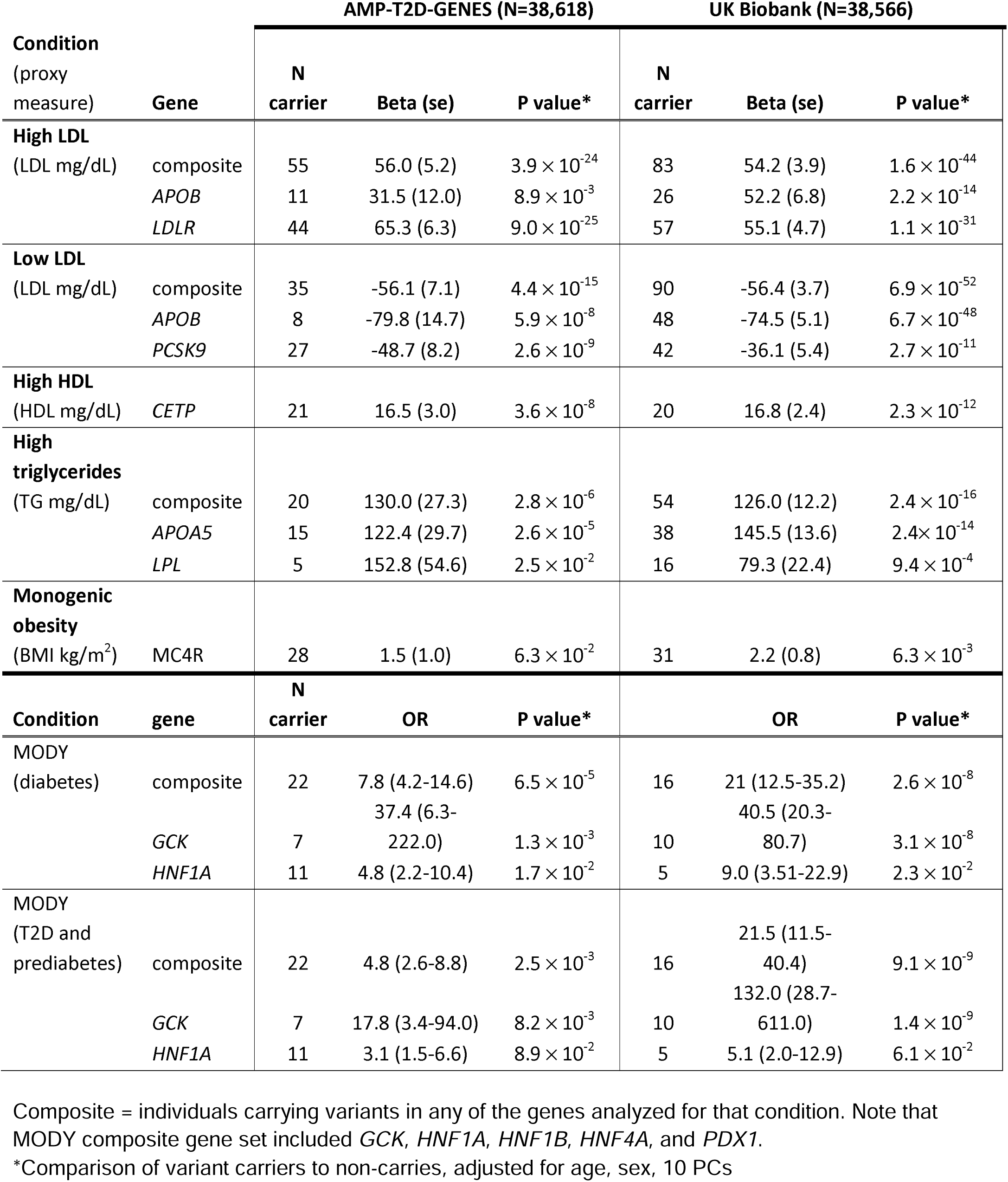
Impact of clinically significant variants on traits.

We also performed the same effect size estimates noted above, but for the variants filtered out during our curation process. We reclassified 7% (21/276) of curated variants from review articles and from ClinVar (which had been designated as clinically significant by at least one submitting source) to “benign” or “likely benign.” Likewise, 27% (59/218) of the pLoF variants were downgraded by our manual review of sequence reads. Together, these ClinVar, review, and pLoF variants which were downgraded by our curation (77/463, 17%) had marked reduced effect sizes compared to variants we curated as clinically significant (**Supplementary Table 5**)^24-27^. These findings support our curation process and highlight the need for caution in relying on available variant designations without additional review.

### Monogenic variant effect sizes are significantly larger than the top 1% of polygenic risk scores

We next directly compared the effect of monogenic variation to common genetic variation on the same corresponding biomarkers in UKB participants. We employed published polygenic scores capturing millions of common genetic variants across the genome, termed global extended polygenic scores (gePS)^28^ (see **Methods**). Since the gePS predicts lifetime risk of developing a disease, and the population mean age in UKB was 58 years, it was possible that estimates by gePS would be under-estimates not capturing individuals who would later in life develop a given condition. We therefore performed gePS analyses restricted to individuals age ≥ 60 year (mean age 65 years) so as to have a fairer comparison with monogenic conditions, which are typically diagnosed at a younger age.

Individuals with the top 1% of gePS had more extreme lipid levels or diabetes risk compared to those with average gePS (25-75%tiles) (**Supplementary Table 6**); however the carriers of clinically significant monogenic variants for these same conditions had even more severe values compared to those top 1% respective gePS’s (*P*<0.05 for each condition, **Figure 2, Supplementary Table 6**). For obesity, the difference in BMI between *MC4R* monogenic variant carriers and the top 1% BMI gePS was not significant (**Figure 2**).

**Figure 2.**
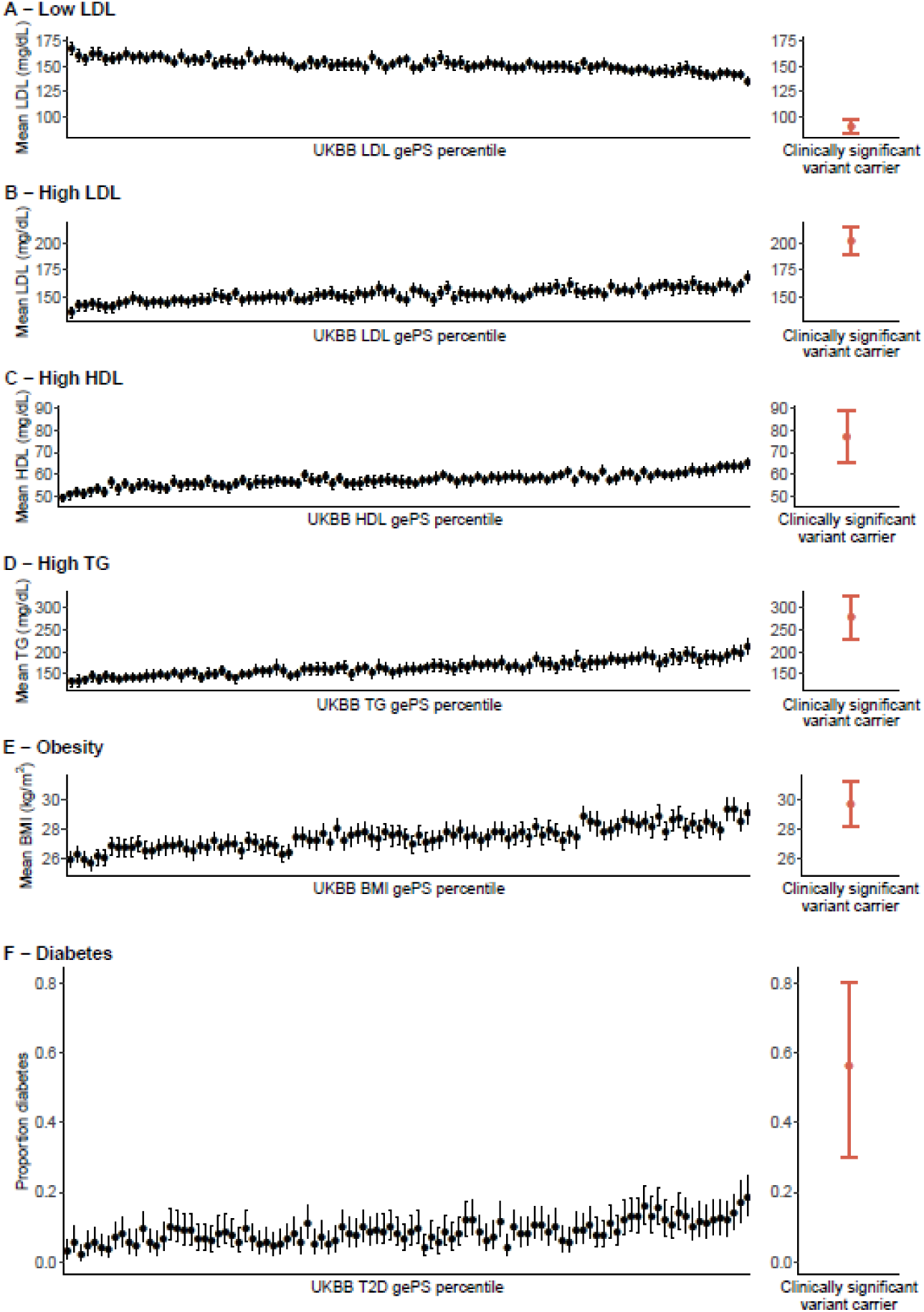
Carriers of rare clinically significant monogenic variants for lipid conditions and monogenic diabetes have more extreme effect size estimates than individuals with the top 1% of global extended polygenic scores (gePS). In all plots, the left panels show the distribution of the phenotype in each percentile of the gePS for the relevant condition (black), and the right panel shows the phenotype distribution in carriers of rare clinically significant monogenic variants for the corresponding condition (red; low LDL cholesterol (*APOB, PCSK9*), high LDL cholesterol (*LDLR, APOB*), high HDL cholesterol (*CETP*), high triglycerides (*APOA5, LPL*), monogenic obesity (*MC4R*), and MODY (*GCK, HNF1A, PDX1*). **A-E)** Mean and 95% CI of each phenotype are indicated by the point and error bars respectively. The same gePS calculated for risk of increasing LDL levels was used for **A** and **B**; however, the inverse of this gePS was used for **B** to illustrate that higher gePS indicates risk of lower LDL cholesterol. **F)** The proportion of individuals with diabetes and 95% CI computed with the Clopper-Pearson method are shown as points and error bars respectively. Individuals in the gePS analysis were restricted to those age ≥ 60 years. LDL cholesterol and triglyceride values were adjusted for lipid-lowering medication use as per methods.

### Monogenic metabolic conditions display highly variable penetrance estimates

While in aggregate clinically significant monogenic variants had marked effect sizes, individual-level trait values in carriers varied considerably (**Figure 3A**). In both datasets, penetrance estimates based on standard disease cut-offs were estimated to be less than 60% for all monogenic metabolic conditions except elevated HDL cholesterol and monogenic diabetes (**Figure 3B, Supplementary Table 2**). Penetrance estimates using composite gene sets for conditions ranged from no more than 20% for low LDL cholesterol conditions caused by *APOB* and *PCSK9* variants (20.0%, 95% CI 8.4-36.9% in AMP-T2D-GENES, 5.6%, 95% CI 1.8-12.5% in UKB) to greater than 80% for diabetes or prediabetes caused by MODY genes *GCK, HNF1A, HNF4A, HNF1B*, and *PDX1* (86.4%, 95% CI’s 65.1-97.1% in AMP-T2D-GENES, 81.2%, 54.4-96% in UKB) (**Figure 3B, Supplementary Table 4**).

**Figure 3.**
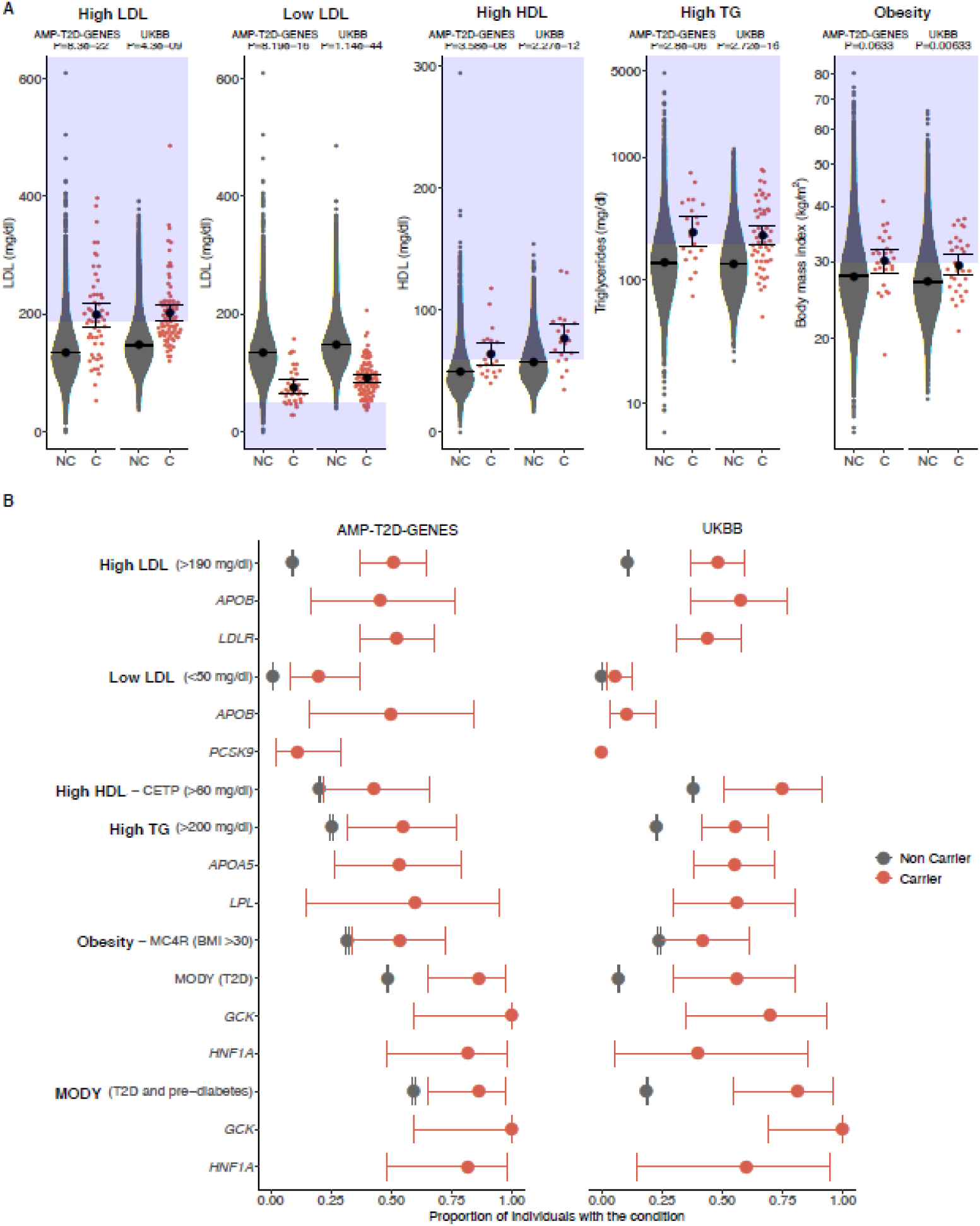
Phenotype distributions and penetrance estimates of clinically significant variant carriers. In all plots, clinically significant variant carriers are shown in red and non-carriers are shown in grey. The left panel of each plot shows AMP-T2D-GENES participants (T2D case/control study) and the right panel shows UK Biobank participants (population-based study). **A)** Mean and 95% CI are represented by the black circle and black lines respectively. Relevant lipid levels (mg/dl) or body mass index (kg/m^2^) are shown for carriers (C) and non-carriers (NC) of clinically significant variants for the five monogenic conditions. The blue boxes indicate the phenotype values that meet a clinical threshold for diagnosis of each of the conditions, and *P* values were obtained by burden analysis in EPACTS (see **Methods**). **B)** Dots are the proportion of individuals that have the condition based on the clinical diagnosis threshold for each condition; for MODY, we show the proportion of individuals meeting T2D as well as T2D and prediabetes criteria (see **Methods**). Error bars reflect 95% CI computed with the Clopper-Pearson method.

### Genetic vs phenotypic ascertainment of MODY suggests broad phenotypic spectrum

We performed deeper phenotyping of MODY variant carriers in the two datasets to determine whether these genetically ascertained individuals manifested clinical features suggestive of MODY, as typically seen in phenotypically ascertained MODY cases. Monogenic diabetes, and particularly MODY (the most common form) can often be misdiagnosed as type or type 2; however, MODY has subtle phenotypic differences from these other forms of diabetes and also, importantly, distinct gene-specific therapeutic strategies^29^.

Focusing on the MODY genes most commonly offered in commercial panels available in the United States (*HNF1A, GCK, HNF4A, HNF1B*, and *PDX1*)^30^, more than 80% of carriers of clinically significant variants had evidence of prediabetes or diabetes (81.2% (95% CI 54.4-96.0%) in UKB and 86.4% (95% CI 65.1-97.1%) in AMP-T2D-GENES) (**Supplementary Table 8, Supplementary Figure 2**). Notably clinical features classically associated with MODY (BMI ≤30 and triglycerides ≤150^31,32^) were only observed in 50% (11/22) of MODY variant-carrying individuals in AMP-T2D-GENES and 75% (12/16) in UKB. Similarly, an expected young age of diagnosis (age ≤35 years), was only observed in 21% (3/14) of those with available data across both datasets (**Supplementary Table 8**). Thus, at least 63% of all MODY variant carriers did not have expected clinical features. Since participants in AMP-T2D-GENES were selected to be T2D cases or controls, and specific exclusion practices employed by several studies to remove possible monogenic diabetes cases (**Supplementary Table 2**)^18^, such ascertainment practices could certainly introduce bias away from classical MODY features.

Nevertheless, when all MODY carriers were compared to others with diabetes in each study, they had significantly lower mean BMI and serum triglycerides (BMI: AMP-T2D-GENES: 26.6 vs 28.7 kg/m^2^, *P*=0.027; UKB: 25.8 vs 31.7 kg/m^2^, *P*=0.004; triglycerides: AMP-T2D-GENES: 136 vs 182 mg/dL, *P*=0.032; UKB: 97 vs 186 mg/dL, *P*=0.004; adjusted for age, sex, and 10 PCs). Thus, in aggregate, MODY variant carriers displayed expected clinical features, but on an individual level, genetically ascertained individuals revealed a broader spectrum of disease phenotype.

We also identified gene-specific findings for the two most common MODY genes, *GCK* and *HNF1A*. GCK-MODY is characterized by non-progressive asymptomatic mild hyperglycemia that is present from birth and may remain in the prediabetes state rather than progress to diabetes^33^. In both datasets 100% (17/17) of carriers of clinically significant *GCK* variants developed diabetes or prediabetes (penetrance estimates of 100%, 95% CI: 59.0-100% in AMP-T2D-GENES and 69.2-100% in UKB). All those with glycated hemoglobin (HbA1c) values available (N=13) had levels consistent with GCK-MODY, ranging from 5.7 to 7.2% (HbA1c in GCK-MODY is typically 5.6-7.6%^34^) (**Supplementary Table 8**). Penetrance estimates for diabetes in HNF1A-MODY from our two datasets (81% in AMP-T2D-GENES, 95% CI 48.2-97.7 and 40% in UKB, 95% CI 5.27-85.3 diagnosed with diabetes by 55 years) were lower than what has previously been reported in the literature (e.g. 97%, 95% CI 96–98 by 50 years^35^) (**Supplementary Table 8**).

### Phenotypic ascertainment strongly impacts estimates of expressivity

It is well-appreciated that phenotypic ascertainment of individuals can upwardly bias estimates of expressivity^14,36^, and we sought to better define this impact by studying conditions of high and low LDL cholesterol levels, where we had information on phenotypic ascertainment within a specific AMP-T2D-GENES cohort. A set of 535 individuals selected for extreme LDL cholesterol (>98^th^ or <2^nd^ percentile), without knowledge of their monogenic condition carrier status, were sequenced as part of the Exome Sequencing Project (ESP) cohort in AMP-T2D-GENES^37^ and not included in the prior analyses. Within this ascertained sample, we identified 18 carriers of clinically significant monogenic high LDL cholesterol variants in *APOB* and *LDLR* (mean LDL 329 mg/dL) and 15 carriers in low LDL cholesterol variants in *APOB* and *PCSK9* (mean LDL 49.2 mg/dL). As expected, compared to carriers of variants for the same LDL cholesterol conditions but not ascertained on LDL phenotype, the two ascertained groups had more extreme LDL cholesterol levels (mean LDL cholesterol values 198 mg/dL, *P*=4×10^−4^ and 77 mg/dL, *P*=0.06 respectively, **Figure 4, Supplementary Table 7**).

**Figure 4.**
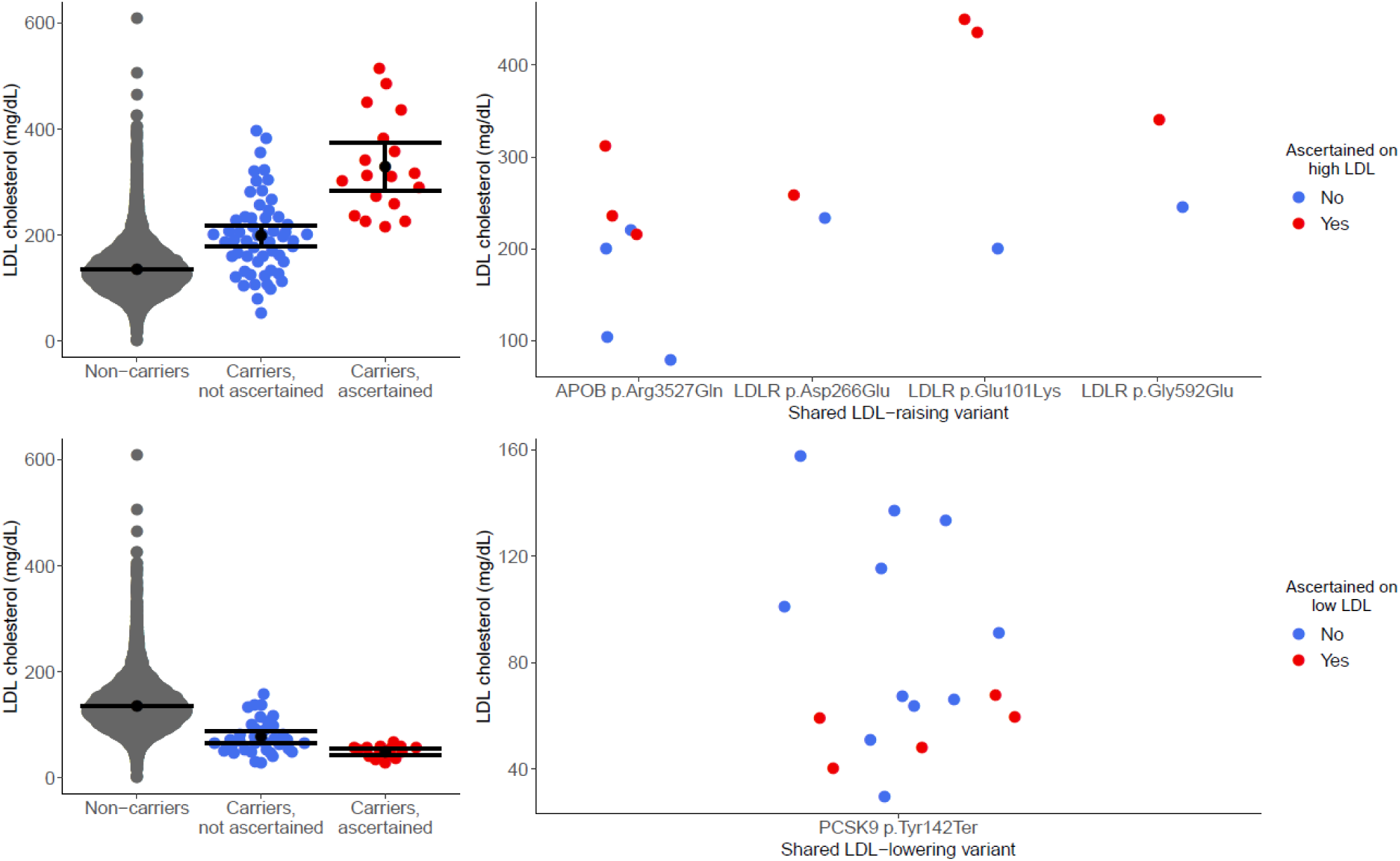
Ascertainment bias significantly impacts expressivity of clinically significant variants for LDL cholesterol conditions. LDL cholesterol levels are shown for carriers and non-carriers of LDL cholesterol raising (top panels) or lowering (bottom panels) clinically significant variants. The variants carriers are stratified by whether they were identified in individuals phenotypically ascertained for extreme serum LDL cholesterol levels (Yes, Red) or in a separate unascertained population (No, Blue) (see **Methods**). The left panels show all clinically significant variant carriers. The right panels show carriers of the single variants that were present in both ascertained and unascertained individuals. LDL cholesterol values are adjusted for lipid-lowering medication use as per methods.

Five variants (High LDL: *LDLR* p.Glu101Lys, *LDLR* p.Asp266Glu, *LDLR* p.Gly592Glu, *APOB* p.Arg3527Gln; Low LDL: *PCSK9* p.Tyr142Ter) were carried by individuals both in the phenotypically ascertained group and in the rest of the AMP-T2D-GENES cohort. These variants showed the same pattern of significantly more extreme LDL cholesterol values in the phenotypically ascertained compared to genetically ascertained individuals (*P*<0.05; all analyses adjusted for age, sex, ancestry, and diabetes status; **Figure 4; Supplementary Table 7**). These marked differences in LDL cholesterol values between the phenotypic vs genetic ascertained carriers, even among those carrying exactly the same LDL cholesterol variant, could not be explained by the use of lipid-lowering medication, assay use, or biased selection of the LDL cholesterol values among those available (e.g. selection of maximum LDL cholesterol value ever for phenotypically ascertained participants)^37^.

In fact, the mean absolute impact of phenotype ascertainment on serum LDL cholesterol levels among individuals with monogenic LDL-raising or lowering variants (27.8-131.0 mg/dL, **Supplementary Table 7, Figure 4**) was thus similar or greater than the mean impact of carrying these same variants compared to non-carriers (31.5-65.3 mg/dL, **Table 1, Figure 4**). Such a substantial effect from phenotypic ascertainment reflects the large variation in expressivity at the single variant level and underscores the importance of considering phenotypic ascertainment bias in monogenic risk prediction.

### Polygenic risk may increase expressivity of monogenic variants

The substantial variability in phenotype expressivity that we observed across all monogenic conditions (**Figure 3A**) suggests that additional environmental and/or genetic factors contribute to expressivity beyond the given monogenic variant. We assessed whether common genetic variation alters expressivity in UKB participants carrying monogenic disease variants.

Among carriers of high HDL cholesterol, low LDL cholesterol, high triglycerides, and monogenic obesity variants, we found that a higher gePS for each condition was associated with a more severe phenotype (e.g., among carriers of monogenic high HDL cholesterol variants, having an increased HDL gePS was associated with even higher HDL cholesterol). However, these trends were only significant for high HDL cholesterol (gePS one SD: beta 17.52 mg/dL, *P*=0.012) and high triglycerides (gePS one SD: beta 80.57 mg/dL, *P*=0.014) (**Figure 5, Supplementary Table 9**). Notably, despite our large study size, power in this analysis was limited, and we estimate that at least 98 carriers of clinically significant variants for a given monogenic condition would be needed for 80% power to detect a correlation of 0.25 (the minimum noted for the above traits) between a given trait and gePS at significance level α=0.05. Therefore, for a monogenic condition with prevalence of 1 in 10,000 individuals, a population-based study with sample size on the order of one million individuals would be required to categorically determine the impact of polygenic risk.

**Figure 5.**
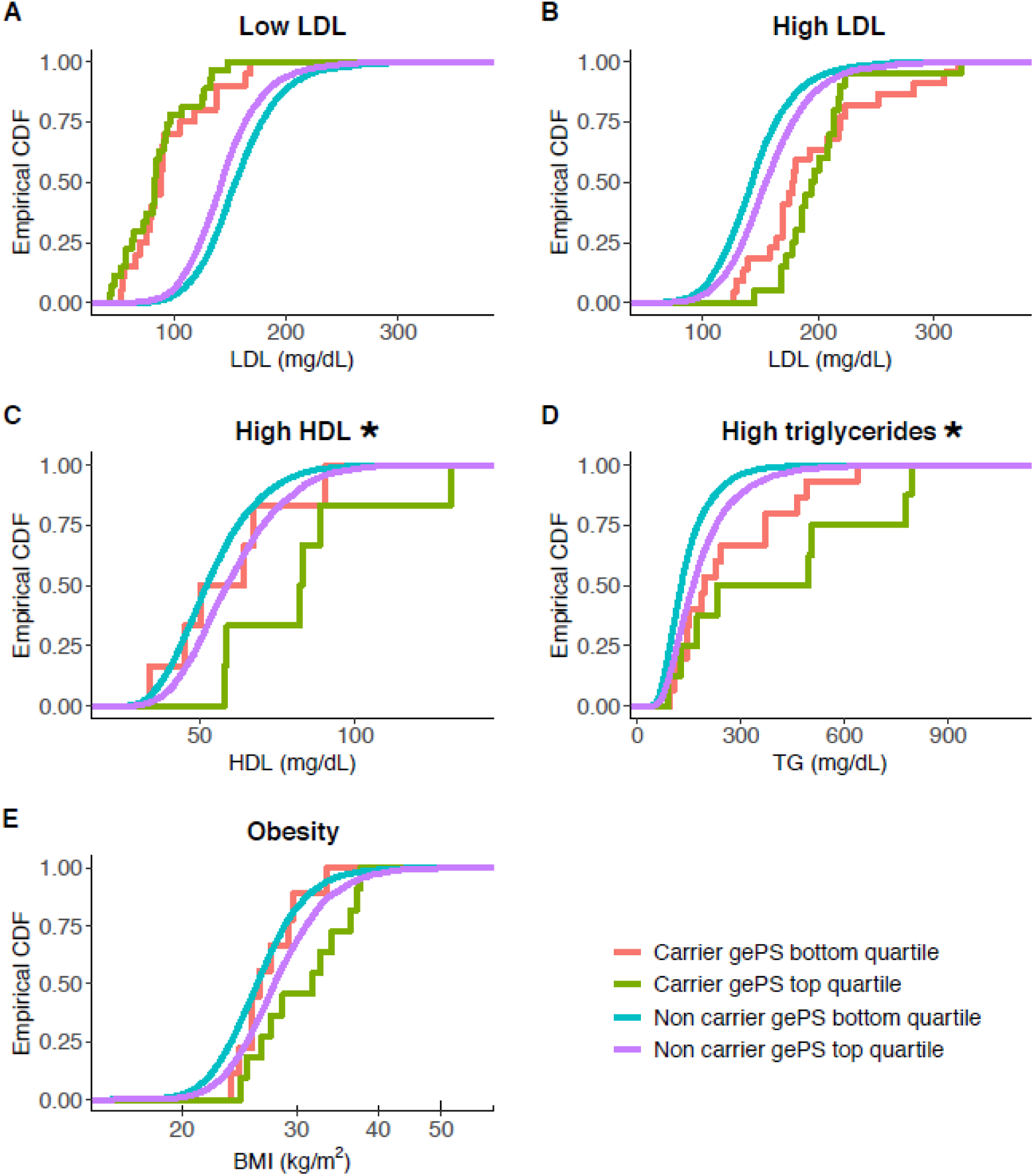
The combination of clinically significant monogenic variants and corresponding polygenic scores significantly improves prediction for high HDL cholesterol and high triglyceride conditions. In all plots, an empirical cumulative distribution function (CDF) of each phenotype is shown for clinically significant variant carriers and non-carriers in the UKB for each monogenic condition stratified by bottom/top quartiles of the corresponding gePS. The monogenic conditions are low LDL cholesterol (*APOB, PCSK9*), high LDL cholesterol (*LDLR, APOB*), high HDL cholesterol (*CETP*), high triglycerides (*APOA5, LPL*), and monogenic obesity (*MC4R*). The same gePS calculated for risk of increasing LDL cholesterol levels was used for **A** and **B**, however the inverse of the gePS was used for **A** to illustrate that higher gePS indicates risk of lower LDL cholesterol. Asterisks indicate *Pvalue*<0.05.

We also assessed the interaction between gePS and monogenic risk in both monogenic carriers and non-carriers in the UKB, and observed significant positive interactions for the same two conditions, high HDL cholesterol (*P*=0.001) and high triglyceride levels (*P*=0.01); however, given the complexities of interaction analyses, additional work will also be needed in larger cohorts before we can conclude that gePS contributes to phenotype expression differently in carriers and non-carriers^38^.

## Discussion

Until recently, the impact of clinically significant monogenic variants on predicting phenotype expression has been predominantly studied in individuals or families ascertained on phenotype^13^. Our analysis employed population-based studies to provide less biased estimates of penetrance and expressivity, and to quantify the impact of phenotypic ascertainment and polygenic risk. We were able to directly compare monogenic and polygenic risk for each condition, and also assess the additional contribution of polygenic risk to expressivity for carriers of monogenic variants.

We applied the current gold standard ACMG/AMP clinical variant classification criteria^9^ to ensure relevance to current clinical practice and demonstrated resultant improvement in risk stratification. Gene variant curation was blinded to participant phenotypes and assessed the pathogenicity of variants expected to cause multiple metabolic conditions in 77,184 exomes of adults (age ≥ 40 years) from the AMP-T2D-GENES consortium and the UK Biobank. Our current analysis adds to a growing set of studies aimed at re-evaluating penetrance estimates using population-based studies^7,9,14-17,39^; however, to the best of our knowledge, this work represents the largest study utilizing clinical standard-of-care ACMG/AMP criteria to curate gene variants in order to establish the penetrance and expressivity of all these monogenic conditions.

Carriers of the highly curated clinically significant variants for MODY and monogenic dyslipidemias had significantly more extreme trait effect sizes compared to non-carriers (OR > 7 for diabetes risk, betas 16.5-130.0 mg/dL for dyslipidemias, Pvalues < 10^−4^, **Table 1**). Despite differences in study populations and designs, the effect estimates for rare monogenic variation for all conditions aside from monogenic diabetes (which was subject to ascertainment bias in AMP-T2D-GENES) were remarkably consistent between the two studies, supporting the integrity of our variant curation. We also assessed the impact of common genetic variation with polygenic scores. There has recently been a great deal of interest around the potential clinical contribution of such scores, especially gePS, and particularly in comparison to monogenic variant risk^6^. We show here that with the exception of monogenic obesity, polygenic risk at the top 1% of the risk distribution is not equivalent to monogenic risk, consistent with recent observations,^7^ but in contrast with others.^40^ There will likely be further development of polygenic scores with improved disease prediction in the coming years^41^; however, in their current state and for the conditions we studied, the risk conferred by polygenic scores on their own was still substantially less than clinically significant monogenic variants; the only exception to this was *MC4R* obesity variants, which are known to have low predictive value for obesity risk^42^.

We observed a wide range of expressivity among clinically significant monogenic variant carriers across all traits (**Figure 2A**), and consequently estimates of penetrance were below 60% for all conditions except elevated HDL cholesterol and monogenic diabetes. At the single gene-level, the penetrance of *MC4R* for obesity (BMI ≥ 30 kg/m^2^) was less than 55%, consistent with previous findings^7,42,43^ (**Figure 2B**), while the penetrance of GCK-MODY was 100% for diabetes or prediabetes (95% CI’s 59-100% in AMP-T2D-GENES, 69-100% in UKB). The range of penetrance estimates across genes and conditions may relate to ability to measure the direct biomarker(s) impacted by a given gene, the extent to which there are redundant mechanisms available in a given pathway to overcome a genetic defect^44^, and the extent to which additional factors, such as other genetic and environmental factors (e.g. diet), impact the trait^12^. The finding of 100% penetrance for diabetes or prediabetes seen in the 17 carriers of GCK*-*MODY across both datasets is particularly intriguing. *GCK* encodes glucokinase, which acts as the cell’s glucose sensor as it facilitates phosphorylation of glucose to glucose-6-phosphate in the pancreatic beta cell, which is the first and rate-limiting step in glucose metabolism^45^. The complete penetrance we have observed may be due to the ability to directly measure glucose as a relevant biomarker, as well as the essential role of *GCK* in glucose homeostasis, with suspected non-redundancy in functioning as a glucose sensor^45^.

We also characterized the impact of phenotypic ascertainment bias on expressivity of clinically significant variants, showing that in individuals with the same LDL cholesterol-raising or -lowering variants there were significant differences in biomarker levels depending on the mode of ascertainment (genetic vs phenotypic) (**Figure 3**) and that the magnitude of this difference on LDL cholesterol levels (29-129 mg/dL) was similar or greater than the mean effect size of such variants (31.5-65.3 mg/dL, **Table 1**). This substantial impact of ascertainment bias was seen at the individual variant level, consistent with other similar observations of LDL cholesterol levels in *LDLR* and *APOB* carriers in a different study population^36,46^ and *HNF4A* p.Arg114Trp in diabetes risk^14^ (*HNF4A* p.Arg114Trp was present in the present datasets, but filtered out due to its designation as a variant of uncertain significant (VUS), reflecting its known low penetrance). The extent of ascertainment bias that we and others have identified highlights an important genetic counseling consideration, particularly with respect to interpretations of genomic sequencing data with limited clinical context available: interpretation of the same test result will likely have different prognostic implications depending on whether the individual tested or family members carry the phenotype of interest (e.g. hyperlipidemia) *vs* if a variant is identified secondarily; a Bayesian framework that takes into account pre-test probability might therefore be useful^47^. Additionally, the variable expressivity seen at the single-variant level in multiple instances further supports additional risk factor modulation from other genetic and environmental exposures.

With regard to additional genetic factors impacting expressivity, we assessed the impact of more common polygenic variation on carriers of monogenic variants and found significant contributions for both high HDL cholesterol and high triglyceride levels (*P*<0.05). These results add to a growing body of research supporting a significant polygenic contribution to monogenic risk^7,39,48,49^. As such analyses were restricted to carriers of monogenic variants, power was limited, and it will be important to investigate in even larger datasets. We estimate that for a monogenic condition with prevalence of 1 in 10,000 individuals, population-based analyses well-powered to capture the contribution of polygenic risk to individuals with the monogenic condition would require on the order of one million individuals.

One limitation of this study is that our selection of variants for curation did not include all possible missense variants, but rather was confined to those reported in ClinVar or subject area reviews. This approach was designed to streamline the variant curation process and restrict our analyses to highly-confident pathogenic variants, but also meant that we were unable to generate estimates of the prevalence of monogenic condition in the two datasets. As discussed previously, there is also the potential for residual bias within the datasets. In the case of AMP-T2D-GENES, ascertainment of participants could have impacted penetrance of monogenic diabetes and expressivity of the metabolic phenotypes (**Supplementary Table 2**). In the UKB, a healthy participant bias^50^ might be expected to reduce estimates of penetrance. Furthermore, despite our large dataset of exomes, the likelihood of observing any specific rare pathogenic variant is still low; this raises the possibility of bias toward lower penetrance of clinically significant variants, since allele frequency is a major predictor of pathogenicity^51^, and rarer variants with potentially greater penetrance are less likely to be observed.

Strengths of this study include the large number of participants with both phenotype and exome data, and the strict variant curation methodology applied. Our analysis of 276 variants designated by ClinVar as pathogenic or likely pathogenic highlights the need for careful curation of variants in clinical practice, with 57% reclassified to “benign,” “likely benign,” or “variant of uncertain significance” with application of ACMG/AMP criteria (**Figure 1**). Of note, however, the ClinVar variants we curated included those submitted to the database before establishment of current standards for curation^9^. With time, we can expect that the ClinVar database will become a more reliable resource for ascertaining clinically significant variants, as more submitters utilize standardized curation practices and additionally as condition-specific standards and curation are provided by ClinGen Expert Panels, including the Monogenic Diabetes Expert Panel in which several of the co-authors participate^52^.

Our study emphasizes the critical need for careful interpretation of monogenic variation, highlighting the roles of variant curation, phenotypic ascertainment, and polygenic risk in the estimates of penetrance and expressivity. In the coming years, access to larger sequencing studies will allow assessment of increasingly rare variants; however, deep phenotyping of such datasets, for example information on medication use and age of disease onset, will to be needed in parallel to better define genetic risk estimates. Improved understanding of monogenic variant expressivity will also likely require broader incorporation of genetic variation across the allelic frequency spectrum and integration of environmental factors. Such advances will facilitate modeling of disease risk and ultimately guide individualized patient genetic counseling and management recommendations.

## Data Availability

Sequence data and phenotypes for this study are either currently available or will be made available via the database of Genotypes and Phenotypes (dbGAP) and/or the European Genome-phenome Archive, as indicated in Supplementary Table 2.

## Acknowledgements

This work was supported by NIH/NIDDK U01 DK105554 to JCF. This research has been conducted using the UK Biobank Resource under application number 27892. MSU is supported by NIH/NIDDK K23 DK114551. AODL was supported by NIH/NICHD K12 HD052896. MB is supported by NIH/NIDDK DK062370. JCF is also supported by NIH/NIDDK K24 DK110550.

Funding for GO ESP was provided by NHLBI grants RC2 HL-103010 (HeartGO), with exome sequencing was performed through NHLBI grants RC2 HL-102925 (BroadGO) and RC2 HL-102926 (SeattleGO). HeartGO components and their support include Atherosclerosis Risk in Communities (NHLBI contracts N01 HC-55015, N01 HC-55016, N01HC-55017, N01 HC-55018, N01 HC-55019, N01 HC-55020, and N01 HC-55021); Cardiovascular Health Study (NHLBI contracts HHSN268201200036C, HHSN268200800007C, N01HC55222, N01HC85079, N01HC85080, N01HC85081, N01HC85082, N01HC85083, and N01HC85086); and NHLBI grants U01HL080295, R01HL087652, R01HL105756, R01HL103612, and R01HL120393, with additional contribution from the National Institute of Neurological Disorders and Stroke. Additional support was provided through R01AG023629 from the National Institute on Aging. A full list of principal Cardiovascular Health Study investigators and institutions can be found at CHS-NHLBI.org; Coronary Artery Risk Development in Young Adults (NHLBI contracts N01-HC95095, N01-HC48047, N01-HC48048, N01-HC48049, and N01-HC48050); Framingham Heart Study (NHLBI contract N01-HC-25195 and grants NS17950, AG08122, and AG033193); Jackson Heart Study (NHLBI contracts N01 HC-95170, N01 HC-95171, and N01 HC-95172); Multi-Ethnic Study of Atherosclerosis (NHLBI contracts N01-HC-95159 through N01-HC-95169 and grant 024156).

Cardiovascular Health Study: This CHS research was supported by NHLBI contracts HHSN268201200036C, HHSN268200800007C, HHSN268201800001C, N01HC55222, N01HC85079, N01HC85080, N01HC85081, N01HC85082, N01HC85083, N01HC85086; and NHLBI grants U01HL080295, R01HL087652, R01HL105756, R01HL103612, R01HL120393, and U01HL130114 with additional contribution from the National Institute of Neurological Disorders and Stroke (NINDS). Additional support was provided through R01AG023629 from the National Institute on Aging (NIA). A full list of principal CHS investigators and institutions can be found at CHS-NHLBI.org. We gratefully acknowledge the Eunice Kennedy National Institute of Child Health and Human Development for support of the MODY variant curation through U24 HD093486 (to TIP). The content is solely the responsibility of the authors and does not necessarily represent the official views of the National Institutes of Health.

The Jackson Heart Study (JHS) is supported and conducted in collaboration with Jackson State University (HHSN268201800013I), Tougaloo College (HHSN268201800014I), the Mississippi State Department of Health (HHSN268201800015I) and the University of Mississippi Medical Center (HHSN268201800010I, HHSN268201800011I and HHSN268201800012I) contracts from the National Heart, Lung, and Blood Institute (NHLBI) and the National Institute on Minority Health and Health Disparities (NIMHD). The authors also wish to thank the staffs and participants of the JHS.

Novo Nordisk Foundation Center for Basic Metabolic Research is an independent Research Center, based at the University of Copenhagen, Denmark and partially funded by an unconditional donation from the Novo Nordisk Foundation (www.cbmr.ku.dk) (Grant number NNF18CC0034900).

The LOLIPOP study is supported by the National Institute for Health Research (NIHR) Comprehensive Biomedical Research Centre Imperial College Healthcare NHS Trust, the NIHR Official Development Assistance (ODA, award 16/136/68), the European Union FP7 (EpiMigrant, 279143) and H2020 programs (iHealth-T2D, 643774). The views expressed are those of the author(s) and not necessarily those of the Imperial College Healthcare NHS Trust, the NHS, the NIHR or the Department of Health. We thank the participants and research staff who made the study possible. JC is supported by the Singapore Ministry of Health’s National Medical Research Council under its Singapore Translational Research Investigator (STaR) Award (NMRC/STaR/0028/2017).

The TwinsUK study was funded by the Wellcome Trust and European Community’s Seventh Framework Programme (FP7/2007-2013). The TwinsUK study also receives support from the National Institute for Health Research (NIHR)-funded BioResource, Clinical Research Facility and Biomedical Research Centre based at Guy’s and St Thomas’ NHS Foundation Trust in partnership with King’s College London.

The views expressed in this article are those of the author(s) and not necessarily those of the NHS, the NIHR, or the Department of Health. MMcC has served on advisory panels for Pfizer, NovoNordisk and Zoe Global, has received honoraria from Merck, Pfizer, Novo Nordisk and Eli Lilly, and research funding from Abbvie, Astra Zeneca, Boehringer Ingelheim, Eli Lilly, Janssen, Merck, NovoNordisk, Pfizer, Roche, Sanofi Aventis, Servier, and Takeda. As of June 2019, MMcC is an employee of Genentech, and a holder of Roche stock. MMcC wishes to acknowledge NIDDK U01-DK105535 and Wellcome: 090532, 098381, 106130, 203141, 212259.

The San Antonio Mexican American Family Studies (SAMAFS) are supported by the following grants/institutes. The San Antonio Family Heart Study (SAFHS) and San Antonio Family Diabetes/Gallbladder Study (SAFDGS) were supported by U01DK085524, R01 HL0113323, P01 HL045222, R01 DK047482 and R01 DK053889. The Veterans Administration Genetic Epidemiology Study (VAGES) study was supported by a Veterans Administration Epidemiologic grant. The Family Investigation of Nephropathy and Diabetes - San Antonio (FIND-SA) study was supported by NIH grant U01DK57295. The SAMAFS research team acknowledges the contributions of late Dr. H. E. Abboud to the research activities of the SAMAFS.

The KARE cohort was supported by grants from Korea Centers for Disease Control and Prevention(4845–301, 4851–302, 4851–307) and intramural grants from the Korea National Institute of Health (2016-NI73001-00, 2019-NG-053-01).

RJFL is supported by the NIH (R01DK110113, R01DK107786, 1R01DK124097). NC is supported by a grant from the Canadian Institutes of Health Research (CIHR Fellowship). The Mount Sinai BioMe Biobank has been supported by The Andrea and Charles Bronfman Philanthropies and in part by Federal funds from the NHLBI and NHGRI (U01HG00638001; U01HG007417; X01HL134588).

The Framingham Heart Study (FHS) acknowledges the support of Contracts NO1-HC-25195, HHSN268201500001I and 75N92019D00031 from the National Heart, Lung and Blood Institute and grant supplement R01 HL092577-06S1 for this research. We also acknowledge the dedication of the FHS study participants without whom this research would not be possible. Dr. Vasan is supported in part by the Evans Medical Foundation and the Jay and Louis Coffman Endowment from the Department of Medicine, Boston University School of Medicine.

## Methods

### Study populations and phenotype curation

#### AMP-T2D-GENES

The complete AMP-T2D-GENES cohort consists of 20,791 cases and 24,440 controls selected from multiple distinct multi-ancestry studies ^18^. The present study includes a subset of 22,875 T2D or prediabetes and 15,743 controls from studies who consented for the data to be used in this analysis, which included Genetics of Type 2 Diabetes (GoT2D), the Exome Sequencing Project (ESP), Lubeck Foundation Centre for Applied Medical Genomics in Personalised Disease Prediction, Prevention and Care (LuCamp), Slim Initiative in Genomic Medicine for the Americas (SIGMA T2D), and T2D-GENES (Type 2 Diabetes Genetic Exploration by Next-generation sequencing in multi-Ethnic Samples). General study characteristics are provided in **Supplementary Tables 1** with more details, including exclusion criteria available in **Supplementary Table 2**, which is adapted from Flannick *et al*., 2019^18^. All samples were approved for use by their home institution’s institutional review board or ethics committee. Analysis of the data was approved by the Mass General Brigham (formerly Partners) institutional review board in Boston, Massachusetts were limited to those participants in each cohort with available DNA who consented to genetic studies.

Phenotype information related to diabetes status was collected by each case-control or cohort study, as previously described in Flannick *et al*. Additionally, we defined prediabetes as any individual with HbA1c ≥ 5.7%, fasting blood glucose ≥ 100 mg/dL, or oral glucose tolerance test (OGTT) 2 hour blood glucose ≥ 140 mg/dL. In individuals who were reported to be on lipid-lowering medication, serum LDL cholesterol and triglyceride levels were adjusted for statin use based on previous studies estimating the impact ^53,54^: we divided LDL by 0.7 and triglycerides by 0.85 as has been previously been implemented^55^. Self-reported ancestry was used, as this was previously shown to correlate well with principal component analysis (PCA) defined ancestry and specific exceptions were dropped from analyses ^18^. Analyses described below used a dataset restricted to individuals in the ‘unrelated analysis set’ (see Flannick *et al*. methods). To provide consistency with the UKB dataset, individuals younger than age 40 were also excluded. Individuals recruited to the Pakistan Genomic Resource cohort were excluded for all analyses involving lipid levels or BMI.

#### UK Biobank

UK Biobank (UKB) is a prospective cohort of approximately 500,000 recruited individuals from the general population aged 40–69 years in 2006–2010 from across the United Kingdom, with genotype, phenotype, and linked healthcare record data ^56^. All participants provided electronic informed consent at their initial visit. Analysis of the data was approved by the Mass General Brigham (formerly Partners) institutional review board in Boston, Massachusetts, and was performed under UK Biobank application 27892.

Direct LDL cholesterol (mmol/L), direct HDL cholesterol (mmol/L), triglyceride (mmol/L), BMI (kg/m2) (field codes: 30780, 30760, 30870, 21001) data were extracted for all individuals. Lipid measurements were converted from mmol/L to mg/dL. The mean for all visits was used in subsequent analyses. The ‘Medication for cholesterol, blood pressure, diabetes, or take exogenous hormones’ fields (6177 and 6153) was used to determine lipid-lowering medication, where an individual was considered to be on lipid-lowering medication if it was recorded at any of the visits. LDL and triglyceride values were adjusted for use of lipid-lowering medication, as described above.

Glycated hemoglobin (HbA1c; field code 30750) was taken as the maximum observed across visits. Since monogenic diabetes may be misdiagnosed as type 1 or type 2 diabetes, we used an inclusive definition of diabetes: possible and probable type 1 or type 2 diabetes was determined in a manner similar to previously described methods ^57^. We also considered individuals as having diabetes if they had ICD10 codes E10-E14 (fields: 41202), and recorded diabetes medication use (fields: 6177, 6153), diabetes ever diagnosed by a doctor (field: 2443), nurse interview codes indicating diabetes (fields: 1220 - any diabetes, 1222 - T1D, 1223 - T2D), or HbA1c ≥ 6.5%. Prediabetes was defined as any individual with HbA1c ≥ 5.7%. We also extracted data for the first recorded age of diabetes diagnosis (fields: 20009, 2976), age, and sex.

This dataset was filtered to only unrelated individuals with European ancestry to facilitate comparisons of biomarkers in analyses using polygenic risk scores. Filtering to unrelated individuals was done using the column ‘used.in.pca.calculation’ in the UKB genotype data sample QC document (ukb_sqc_v2.txt) as a proxy. This column indicates samples which UKB used in a principal component analysis (PCA), and this analysis was only performed on unrelated, high quality samples. To filter to European ancestry only, samples were first projected onto 1000 Genomes phase 3 ^58^ PCA coordinate space. Then Aberrant R package ^59^ clustering was used to identify individuals falling within 1000 Genomes project EUR PC1 and PC2 limits (lambda=4.5). Individuals that self-reported as non-European ethnicity were also filtered. There were 38,566 individuals remaining after all filtering and intersection with individuals that also have exome sequence data released in the first tranche (Category 170).

### Generation of gene list

We sought to include genes that would be ordered in the United States in clinical practice to diagnose conditions of monogenic diabetes, lipodystrophy, obesity, and lipid disorders. We searched the Genetic Testing Registry (https://www.ncbi.nlm.nih.gov/gtr/) and Concert Genetics (https://app.concertgenetics.com/), last accessed March 14^th^, 2018, for lists of available commercial gene panels for clinical genetic testing for these diseases available in the United States. We filtered this list of genes to those with an autosomal dominant mode of inheritance, as determined by the Online Mendelian Inheritance in Man® (OMIM, https://www.omim.org/). For the genes in OMIM where mode of inheritance was not specified, the genes were researched in ClinVar (https://www.ncbi.nlm.nih.gov/clinvar/) and related literature. In total there were 26 autosomal dominant genes across the conditions. We further excluded any gene where there was no ClinVar submission (April 2019 ClinVar submission summary) of pathogenic or likely pathogenic for the phenotype of interest that also included clinical testing as a collection method, leaving 20 genes. We determined phenotype overlap by manual review of the “SubmittedPhenotypeInfo” and “ReportedPhenotypeInfo” fields in the submission summary where present and ‘ExplanationOfInterpretation’ or submitted PubMed articles when phenotype info was not reported in the other fields. For MODY, most commercial panels available in March, 2018 included *HNF1A, HNF4A, GCK, HNF1B*, and *PDX1*, with larger panels less widely available. We therefore separated the MODY genes into two categories: “MODY” including those five genes and “MODY extended” including eight additional genes.

### Determination of genes with LoF mechanism

The pLoF curation was restricted to genes alleged to cause disease with a LoF mechanism based on reporting in ClinVar or a PubMed publication of an LoF variant in an individual with the phenotype of interest.

Two genes were determined to be related to both high LDL (familial hypercholesterolemia) and low LDL (familial hypobetalipoproteinemia): *APOB* and *PCSK9*. Gain-of-function missense mutations in both genes result in increased LDL levels, while LoF mutations cause lower LDL levels ^60-62^. Therefore, only missense ClinVar variants in *APOB* and *PCSK9* were assessed in the curation process for high LDL, and LoF variants were considered for low LDL.

### Exome data variant filtering and annotation

All filtering and annotation described below was performed using Hail 0.2 (https://hail.is).

#### AMP-T2D-GENES

Exome sequencing and quality control were described previously ^18^. We applied additional genotype filters to retain only high-quality genotypes: genotype quality ≥ 20, depth ≥ 10, and minor allele balance > 0.25 for heterozygous genotypes. Variants were annotated using Ensembl’s Variant Effect Predictor (VEP) v85 ^63^ with the Loss-of-function Transcript Effect Estimator (LOFTEE) plugin ^64^. The dataset was then filtered to only variants with a consequence on any of the genes of interest. The filtered VCF was used in analyses described below that involve EPACTS.

We determined which variants in our dataset have been submitted to ClinVar by cross-referencing this filtered variant list with the ClinVar VCF (April 2019) (further curation described below). A list of predicted loss-of-function (pLoF) variants, including stop gained, frameshift or essential splice site (splice donor or splice acceptor), was generated by filtering to variants with a LOFTEE high-confidence (HC) annotation on any transcript. Finally, we used transcript expression-aware annotation ^65^ to add pext (proportion expression across transcripts) values for the worst consequence annotation to each variant for use in pLoF curation discussed below.

#### UK Biobank

UKB exome sequencing PLINK files were imported into Hail and all the same annotation described for AMP-T2D-GENES was added using appropriate files for genotype reference GRCh38 and VEP v95. In order to compare UKB variants to AMP-T2D-GENES variants we used Hail’s liftover method to lift data from GRCh38 to GRCh37. Since the PLINK files do not contain genotype quality information that we can use for filtering low-quality genotypes, we downloaded the gVCFs for all variant carriers and determined which individuals genotypes were not high-quality (genotype quality ≥ 20, depth ≥ 10, and minor allele balance > 0.25 for heterozygous genotypes) and set each of these to missing in the VCF.

### ClinVar variant curation

We identified individuals carrying variants in the genes of interest that had at least one ‘pathogenic’ or ‘likely pathogenic’ submission in ClinVar by a clinical testing lab for the relevant trait. To streamline variant curation we first generated a list of high confidence clinical genetic testing laboratories. Using the April 2019 release of the ClinVar submission summary, a lab was considered high confidence if it had submitted more than 15,000 variants to ClinVar and had updated its submission after 2017 when the most recent ACMG variant interpretation guidelines were published ^9^. This resulted in eight labs: Invitae; GeneDx; Ambry Genetics; EGL Genetic Diagnostics; Eurofins Clinical Diagnostics; PreventionGenetics; Laboratory of Molecular Medicine, Partners Healthcare Personalized Medicine; Genetic Services Laboratory, University of Chicago; and Counsyl. Variants that were reported by any lab on this list since January 1st, 2017 were then accepted as having the pathogenicity reported by the lab.

These labs were further verified through manual curation. First, five variants from each lab that were also present in our study were chosen to be manually curated, so that the manual curation could be compared to the lab’s analysis. Through this, we found no differences in curation results. Then, five variants from each lab were chosen at random through ClinVar – one Pathogenic, one Likely Pathogenic, one VUS, one Likely Benign, and one Benign. As PreventionGenetics only submitted Benign and Likely Benign to ClinVar, their variants were limited to those categories. These variants were then also manually curated, and the results were compared. The only difference in curation of the non-study variants involved University of Chicago, due to internal data initially not available to our study curator; however, the same conclusion was reached upon inclusion of this internal data, which was included in their reporting in ClinVar. During the manual phenotype curation (described below), we discovered Counsyl reported conflicting phenotypes for the same variant, so we opted to manually curate variants assessed by Counsyl.

The variants not analyzed by high confidence labs were analyzed separately using manual curation with the curator blinded to carrier phenotypes. The ClinGen Variant Curation Interface (https://curation.clinicalgenome.org/) was used to analyze the variants and assign evidence following the ACMG guidelines ^9^ and recommendation for interpretation of LoF variants^66^, with input from gene-specific rules under development by the Monogenic Diabetes Expert Panel VCEP (https://clinicalgenome.org/affiliation/50016/) for the MODY variants. Databases and other resources such as ClinVar (https://www.ncbi.nlm.nih.gov/clinvar/), Human Gene Mutation Database (HGMD) (https://digitalinsights.qiagen.com/products-overview/clinical-insights-portfolio/human-gene-mutation-database/), gnomAD (https://gnomad.broadinstitute.org/), PubMed (https://pubmed.ncbi.nlm.nih.gov/), Google Scholar (https://scholar.google.com/), Alamut (https://www.interactive-biosoftware.com/alamut-visual/), and the UCSC browser (https://genome.ucsc.edu/) were utilized to collect evidence for curation purposes. The general guidelines were adjusted slightly for certain criteria such as control population frequency as shown in **Supplementary Table 10**. Since most AMP-T2D-GENES participants are included in gnomAD, AMP-T2D-GENES allele frequency decisions were made by subtracting the number of AMP-T2D-GENES carriers from the number of total gnomAD carriers to determine an adjusted gnomAD allele frequency which was compared to the cut-offs shown in **Supplementary Table 10**.

Three variants within *HNF1A* were excluded from further analysis because of poor genotyping quality at this site making it difficult to determine which individuals are actually carriers (GRCh37: 12-121432114-CG-C, 12-121432116-G-GC, 12-121432117-G-GC, GRCh38: chr12:120994311-CG-C, chr12:120994313-G-GC, chr12:120994314-G-GC). As all three are frameshifts, these variants were also excluded from the pLoF curation described below.

Variants in MODY genes were curated by a second set of reviewers at University of Maryland School of Medicine, the home institution of the ClinGen Monogenic Diabetes Expert Panel, to ensure accuracy. All variants were consistently classified as collectively pathogenic or likely pathogenic (**Supplementary Table 12**).

All variants curated for this project, along with their classification and supporting evidence, were submitted to ClinVar on January 30th, 2020.

### High confidence Loss of Function variants

As described above, we used LOFTEE^64^ to generate a list of high confidence pLoF variants, restricting to the set of genes we determined to have a LoF mechanism of pathogenicity. Each pLoF variant was assessed by manual review of reads by two independent reviewers. The reads were examined for poor quality, homopolymer artifacts, and multinucleotide variants (MNVs) causing a synonymous or missense variant instead of the reported stop codon. Where available, gnomAD data was examined to identify variants that were flagged as filtered by gnomAD’s random forest variant quality control method. UCSC genome browser data was assessed to determine the conservation of the region, the location of the variant, and how many transcripts the variant was coding. If the variant was present in the last exon or last 50 base pairs of the penultimate exon, it was deemed not LoF due to a predicted lack of nonsense mediated decay. However, this was overruled if the variant was predicted to delete over 25% of the gene. The potential for a splice site rescue was assessed by examining +/- 21bp around the variant. Any inframe splice site within 6bp was considered an essential splice site rescue and possible inframe splice site rescues between 6 and 21bp were considered a rescue if validated by the alternative splice site prediction tool Alamut (v.2.11). We also used pext values obtained from the transcript expression-aware annotation^65^ to indicate variants that fell in exons that have evidence of poor expression (specific cutoffs are detailed in **Supplementary Table 11**). Variants were classified into 5 categories, ‘LoF’, ‘likely LoF’, ‘uncertain’, ‘likely not LoF’, or ‘not LoF’ using the guidelines described in **Supplementary Table 11**. Any variant that had a discordant assessment between the two reviewers (‘LoF’ or ‘likely LoF’ by only one reviewer) was examined by a third reviewer to determine the final pLoF annotation.

### Carrier vs non-carrier effect size analysis

We considered an individual to be a carrier of a clinically significant variant if they carry a ClinVar variant assessed as pathogenic or likely pathogenic or a pLoF variant passing manual curation (‘LoF’ or ‘likely LoF’ as described above). For AMP-T2D-GENES, as previously described ^18^, we accounted for the diverse ancestry and different sequencing technologies by using a modified version of EPACTS (http://genome.sph.umich.edu/wiki/EPACTS) that sets specified variants to missing based on QC of sample subgroups (as described in Flannick et al, there are 25 subgroups that were determined by stratifying samples by cohort of origin, ancestry, and/or sequencing technology). As covariates in AMP-T2D-GENES analyses, we included sex, age, PCs 1-10, sample subgroup, and sequencing technology all as previously defined ^18^. Analyses on UKB used covariates for sex, age, PCs 1-10 and the genotyping array.

For both AMP-T2D-GENES and UKB, we used VCFs produced after filtering variants as described above and performed the group b.burdenFirth for binary traits and q.burdentest for continuous traits in EPACTS to compare carriers and non carriers for the following condition/phenotype pairs: high LDL cholesterol with LDL cholesterol (mg/dL); low LDL cholesterol with LDL cholesterol (mg/dL); high HDL cholesterol with HDL cholesterol (mg/dL); high triglycerides with triglycerides (mg/dL); monogenic obesity with BMI (kg/m^2^), MODY with diabetes status, and in diabetes cases only: HDL cholesterol, Triglycerides, and BMI.

Additionally, we included T2D or T2D with prediabetes as covariates in all tests on lipid measurements and BMI. Triglycerides and BMI were log transformed. All of these analyses were also performed per gene to ensure that we captured possible gene level differences in phenotype values.

### Estimation of penetrance

Unlike diabetes, phenotypes used to assess the possibility that individuals have each lipid condition or obesity, are continuous. There are commonly used clinical guidelines for diagnosis of each of these conditions, so these cutoffs were used to dichotomize the phenotypes allowing us to determine individuals with and without each condition for use in estimating penetrance. The following clinical diagnosis cutoffs were used: High LDL cholesterol: LDL cholesterol ≥190 mg/dL, Low LDL cholesterol: LDL cholesterol ≤50 mg/dL, High HDL cholesterol: HDL cholesterol ≥60 mg/dL, High triglycerides: triglycerides ≥200 mg/dL^67^, and Monogenic obesity: BMI ≥ 30 kg/m^2^.

Penetrance estimates were calculated as the proportion of individuals carrying a clinically significant variant that also exhibit the expected condition. To determine the significance for all penetrance estimates we used the group Firth burden test in the modified version of EPACTS and the same covariates as described in ‘Carrier vs non-carrier enrichment analysis’.

### Calculation of global extended polygenic score (gePS)

#### Body mass index and type 2 diabetes

Global extended polygenic scores for T2D and BMI were previously calculated on UKB participants using LDpred^6,43^. The variants and weights used in the calculation were downloaded (http://www.broadcvdi.org/informational/data). These weights were then applied to the UKB genotype data from the subset of individuals included in this study to calculate a gePS using Hail’s equivalent to the --score method in PLINK (https://hail.is/docs/0.2/guides/genetics.html?highlight=prs). These values were then scaled and centered around zero with a standard deviation of one for downstream analysis. We confirmed that plots of T2D prevalence and BMI by respective polygenic scores converged at the same upper limits as previously published ^6,43^.

#### Lipid conditions

To estimate a gePS for each lipid phenotype, we filtered UK Biobank genotype data to only the individuals used in this study (unrelated, EUR ancestry, and exome sequenced) and excluded SNPs with an imputation INFO <0.3 and allele frequency <1%. Summary statistics for lipid GWAS were downloaded from the European Network for Genetic and Genomic Epidemiology (ENGAGE) Consortium. This included LDL cholesterol, HDL cholesterol, and triglyceride GWAS summary stats from a meta-analysis of up to 62,166 individuals of European ancestry ^68^. We filtered to variants observed in HapMap3 (--only-hm3) and both the summary statistics and genotype data, and then estimated SNP weights using the Bayesian computational method LDpred (version 1.0.6) which accounts for local LD patterns ^69^. SNP weight estimates were obtained using the infinitesimal (inf) model (assumes all genetic variants impact phenotype) with heritability estimates (TG: 0.1525, LDL: 0.1347, HDL: 0.1572) as previously calculated using LD Score regression ^70^ and displayed on LD Hub ^71^. We then used PLINK version 1.9 (--score) to calculate polygenic scores using the SNP weights ^72^.

As in the BMI and T2D gePRS, the distribution was scaled to have a mean of zero and one standard deviation around the mean. Since there is a single gePS for LDL cholesterol, the scaled gePS was multiplied by -1 for figures and analyses comparing low LDL cholesterol carrier phenotype values to phenotypes aggregated by gePS deciles or quantiles.

#### Statistical Analysis

We used generalized linear models (GLM) to examine the gePS results in a few different ways. We compared the top 1% to the interquartile range (25-75%) of the gePS and to the clinically significant variant carriers (**Supplementary Table 6**). For both analyses we restricted the age in controls to >=60. Additionally, we determine the effect size of gePS on phenotypes in the subset of only clinically significant variant carriers and assessed the interaction of carrier status and gePS (**Supplementary Table 9**). In all GLMs age, sex and 10 PC’s were included in the model as covariates. A linear regression was performed for all phenotypes except diabetes where a logistic regression was applied.

All plots were made using R version 3.5.2.

## Data availability

Sequence data and phenotypes for this study are available via the database of Genotypes and Phenotypes (dbGAP) and/or the European Genome-phenome Archive, as indicated in Supplementary Table 2.

## Code availability

Code used in analyses is will be made available prior to publication in GitHub.

## List of Supplementary Tables

Supplementary Table 1 - Summary characteristics of study populations.

Supplementary Table 2 - Detailed characteristics of study cohorts. AMP-T2D-GENES Cohort Information.

Supplementary Table 3 - Counts of clinically significant variants and carriers across conditions.

Supplementary Table 4 - Effect size and penetrance estimates of clinically significant monogenic variants across conditions and by gene.

Supplementary Table 5 - Effect size and penetrance in filtered out variants (designated benign, likely benign, or not pLOF)

Supplementary Table 6 – Comparison of Top 1% gePS with interquartile range and monogenic carriers.

Supplementary Table 7 - Mean serum LDL values based on ascertainment.

Supplementary Table 8 - Clinical characteristics of monogenic diabetes variant carriers.

Supplementary Table 9 - Impact of polygenic score on trait expressivity in monogenic carriers.

Supplementary Table 10 - Frequency cut-offs used for ClinVar variant curation.

Supplementary Table 11 - pLoF curation categories and guidelines for classification.

Supplementary Table 12 - Variant curation assessments and carrier counts.

## List of Supplementary Figures

Supplementary Figure 1. Distribution of clinically significant variants across ancestries.

Supplementary Figure 2. Carriers of clinically significant variants in MODY genes show a younger age of diabetes diagnosis compared the rest of the AMP-T2D-GENES cohorts and the UK Biobank population.

## Notes

### Competing Interest Statement

The authors have declared no competing interest.

### Author Declarations

All samples were approved for use by their home institution's institutional review board or ethics committee. Analysis of the data was approved by the Mass General Brigham (formerly Partners) institutional review board in Boston Massachusetts.

